# APOE Isoform-Dependent Self-Association Measured by a Split-Luciferase Complementation Assay: Differential Effects of Disease-Risk and Protective Variants

**DOI:** 10.64898/2026.05.09.26352797

**Authors:** Daria Andrieieva, Isabella Falltrick, Cho-Ying Chiang, Bobby Beaumont, Yann Le Guen, Changlu Liu, Siria Pergolesi, Chen-Ting Ma, Michael R. Jackson, Bradley T. Hyman, Rosemary J. Jackson

## Abstract

Apolipoprotein E (ApoE) is the principal lipid transport protein in the central nervous system and the strongest genetic modifier of late-onset Alzheimer’s disease (AD) risk. The three common isoforms, ApoE2, ApoE3, and ApoE4, differ in their propensity to self-associate, with ApoE4 forming oligomers more readily than ApoE3 or ApoE2. This enhanced self-association is proposed to reduce the pool of lipid-competent monomeric ApoE4 available for cholesterol transport and amyloid-β clearance, contributing to AD pathogenesis. Here we describe a quantitative, cell-based split-luciferase complementation assay for ApoE self-association using the NanoBiT system, in which SmBiT- and LgBiT-tagged ApoE produced by HEK293 cells are combined and luminescence is measured. ApoE4 shows significantly enhanced self-association relative to ApoE3, while ApoE2 is no different from ApoE3. Testing a panel of naturally occurring and engineered variants demonstrates that the C-terminal self-association interface is the primary determinant of isoform-specific differences: two *APOE* ε3-backbone C-terminal variants, Jacksonville (V236E) and W276C, both reduce self-association below ApoE3 levels, while the *APOE* ε*4*-backbone protective variant R251G and the engineered domain-interaction probe R61T both reduce ApoE4 self-association to the level of ApoE3. In contrast, the Christchurch variant (R136S), the African-ancestry risk variant R145C, and the Admixed American risk variant R189C do not alter self-association. These findings establish a sensitive cell-based assay for ApoE self-association and highlight the C-terminal domain as a potential therapeutic target for normalizing ApoE4 function.

## INTRODUCTION

The Apolipoprotein E (*APOE*) ε4 allele is the strongest genetic risk factor for late-onset Alzheimer’s disease (AD), increasing risk three-to four-fold in heterozygotes and more than ten-fold in homozygotes, while *APOE*-ε2 is protective and *APOE*-ε3 is risk-neutral(1–3). *APOE* encodes apolipoprotein E (ApoE), a 34 kDa glycoprotein secreted by astrocytes and microglia that packages with cholesterol and other lipids into lipoprotein(4, 5). The three isoforms differ at two residues: ApoE2 carries Cys112/Cys158, ApoE3 carries Cys112/Arg158, and ApoE4 carries Arg112/Arg158. These substitutions produce consequential differences in protein structure and function that translate into markedly divergent effects on AD risk(6, 7).

ApoE comprises an N-terminal receptor-binding domain (residues 1–191) and a C-terminal lipid-binding domain (residues 210–299) separated by a hinge domain (8–10). Although the tendency to self-association has precluded most attempts at a full length structure of ApoE it is predicted that in ApoE4, the Arg112 substitution promotes an intramolecular ‘domain interaction’ between Arg61 and Glu255 that repositions the C-terminal domain, reducing lipid-binding efficiency and impairing intracellular trafficking (11). All three isoforms form higher-order oligomers in lipid-free conditions with species ranging from dimers to assemblies of four to six molecules, and ApoE-containing dimers have been detected in cerebrospinal fluid, frontal cortex, and hippocampus(12–15). Whether the monomer–oligomer equilibrium is functionally significant, for example by regulating the pool of lipid-competent ApoE available for cholesterol transport and amyloid-β clearance, remains an open question (15–17). However only the monomer of ApoE is able to bind lipids and lipidated ApoE appears to be protected from aggregation highlighting this as an important element of ApoE biology for further study(17, 18).

Several rare *APOE* variants modulate AD risk and provide natural experiments for probing the functional importance of self-association. The *APOE3*-backbone Jacksonville variant (V236E), located in the C-terminal coiled-coil interface, reduces ApoE self-aggregation and enhances lipidation, and carriers have AD risk equivalent to *APOE-*ε2 homozygotes(15, 19). Similarly, the recently identified variant W276C was discovered in the *APOE-ε3* backbone to be protective (20). The *APOE-ε4*-backbone variant R251G, also located within the C-terminal domain, counterbalances the AD risk conferred by ε4 when co-inherited(19). In contrast, the protective Christchurch variant (*APOE3\**R136S)(21), the detrimental AD-risk increasing *APOE3\**R145C substitution when combined with *APOE*-ε4 (22) and a second recently discovered variant in *APOE3*\*R189C (OR = 3.35, increased risk)(20), are all in the N-terminal domain.

Existing approaches for measuring ApoE self-association, including size-exclusion chromatography, analytical ultracentrifugation, and native PAGE, are low-throughput and require large quantities of purified protein(8, 23–25). The NanoLuc® Binary Technology (NanoBiT; Promega) splits NanoLuc luciferase into two subunits, Large BiT (LgBiT, 18 kDa) and Small BiT (SmBiT, 1.3 kDa), that associate with low intrinsic affinity (Kd ∼ 190 μM), producing luminescence only when the tagged proteins interact(26). NanoBiT has been applied to secreted proteins and used to study dimerization of SOD1, GPCRs, RAF kinases, and viral proteases(27–29). Here, we adapt the NanoBiT system to measure ApoE self-association in conditioned medium from transfected HEK293 cells and use it to systematically characterize common isoforms, naturally occurring risk and protective variants, and an engineered structural probe. We show that self-association is selectively altered by variants mapping to the C-terminal oligomerization interface, identifying this region as a potential therapeutic target for correcting ApoE4 function.

## MATERIALS AND METHODS

### Plasmid Construction

Full-length ApoE2, ApoE3, and ApoE4 were cloned into expression vectors containing the SmBiT or LgBiT tags of the NanoLuc Binary Technology (NanoBiT; Promega) system, positioned at the C-terminal domain of each isoform. Plasmid DNA was amplified using the QIAGEN Plasmid Maxi Kit (Qiagen, Germany) according to the manufacturer’s instructions. Sequence integrity was confirmed by Sanger sequencing performed by the MRC DNA Core (University of Dundee).

Missense variants were introduced into the appropriate ApoE backbone by site-directed mutagenesis using the Q5® Site-Directed Mutagenesis Kit (New England Biolabs) following the manufacturer’s protocol. Mutagenic primers were designed using the NEBaseChanger® Primer Design Tool (New England Biolabs) and primers are shown in supplemental table 1. Mutant plasmids were isolated using the QIAGEN Plasmid Mini Kit (Qiagen, Germany) and confirmed by Sanger sequencing.

### Cell Culture and Transfection

Human Embryonic Kidney 293 (HEK293) cells were maintained in Dulbecco’s Modified Eagle’s Medium (DMEM) supplemented with 10% (v/v) foetal bovine serum (FBS), 1% (v/v) penicillin-streptomycin (P/S), and 1% (v/v) GlutaMAX™ (Gibco). Cells were passaged at 80% confluency in T75 flasks at a 1:10 split ratio and incubated at 37 °C in 5% CO2.

For transfection, HEK293 cells were seeded at 0.1 × 106 cells per well in 12-well plates (Nunc™ Non-Treated Multidishes) in 1 ml normal media. After 24 hours, cells were transfected using Lipofectamine-2000 (Invitrogen, UK) at a cDNA:lipofectamine ratio of 1:1 (v/v) according to the manufacturer’s instructions. Media was replaced 4–6 hours post-transfection with either Gibco Opti-MEM™ Reduced Serum Media. Cells were incubated for a further 48 hours at 37 °C. Conditioned medium was collected and centrifuged at 10,000 rpm for 3 minutes to remove cell debris, then stored at −20 °C.

### Western Blot

To normalize ApoE concentrations across samples, 13 μl of conditioned medium was combined with 5 μl NuPAGE™ LDS Sample Buffer (4×) and 2 μl NuPAGE™ Sample Reducing Agent (10×) (both Invitrogen™) and heated to 75 °C for 10 minutes. Proteins were resolved on NuPAGE Bis-Tris 4–12% gels in 1× NuPAGE™ MES SDS Running Buffer at 120 V for approximately 1 hour, using 5 μl SeeBlue™ Plus2 Pre-stained Protein Standard (Invitrogen™) as a molecular weight marker. Proteins were transferred to nitrocellulose membranes using the Invitrogen™ iBlot™ 2 Transfer Stack system (7 minutes, 20 V). Membranes were blocked in 5% (w/v) skimmed milk in 1× TBST for 1 hour at room temperature, then incubated overnight at 4°C with anti-ApoE primary antibody (Novus Biologicals/Bio-Techne, catalogue #NBP1-31123; 1:2,000 dilution in blocking solution). After washing, membranes were incubated with IRDye® 800CW Donkey anti-Rabbit secondary antibody (LI-COR Biosciences, catalogue #926-32213; 1:5,000 in 1× TBS) for approximately 1 hour. Membranes were imaged on an Odyssey® CLx Imaging System (LI-COR Biosciences) and band intensities quantified using Image Studio™ 6.0 (LI-COR Biosciences). ApoE concentrations were normalized to the lowest signal within each experimental batch.

### Self-association Assay

Following western blot-based normalization of ApoE concentrations to the lowest signal in each batch, 20μl of ApoE-SmBiT conditioned medium and 20μl of ApoE-LgBiT conditioned medium were combined in white 96-well plates. Following normalization, this equal-volume combination resulted in an approximate 7:1 SmBiT:LgBiT protein ratio, ensuring that ApoE-LgBiT was the rate-limiting component. The NanoBiT substrate solution was prepared by diluting 0.4 μl HiBiT Lytic Substrate in 59.5 μl of assay buffer (0.25% Pluronic® acid, 20 μM HEPES in HBSS [+/+]) per well and added to each sample. Luminescence was measured using a GloMax® Navigator Microplate Luminometer (Promega) at one-minute intervals for 45 minutes. Maximum relative light unit (RLU) values were used for quantification.

To control for the amount of ApoE-LgBiT present in each well, a HiBiT loading control was performed following the self-association read. A HiBiT peptide solution (0.1 μg in 10 μl of assay buffer per well) was added to each well, the plate was equilibrated for 15 minutes, and a single luminescence reading was taken. All conditioned medium was collected in phenol red free media to eliminate background luminescence. As samples were normalized to equal relative ApoE input across constructs on the basis of western blot band intensity, absolute ApoE concentrations were not determined. Accordingly, the assay provides a comparative self-association readout across matched relative-input conditions rather than a direct measure of binding affinity.

### Size-Exclusion Chromatography

Size-exclusion chromatography (SEC) was performed on conditioned medium using a Superdex 200 Increase 10/300 GL column (Cytiva) driven by an ÄKTA Purifier UPC 100. For complementation experiments, equal volumes (0.5 ml) of ApoE-SmBiT and ApoE-LgBiT conditioned medium were combined following western blot-based concentration normalization and loaded onto the column. Fractions of 0.25 ml were collected across the elution profile. Luciferase activity was measured in each fraction by addition of NanoGlo substrate (Promega) in 25 mM Tris-HCl, 150 mM NaCl, pH 7.4 with 0.05% antifoam, and luminescence was read on a FlexStation 3 plate reader (Molecular Devices). Fraction luminescence was normalized to the total signal across all fractions within the elution range expected to contain ApoE, to determine the percentage of total signal attributable to each oligomeric species. To quantify the distribution of ApoE-LgBiT independently of complementation, 30 ng of HiBiT reagent per well and 0.05 μl of NanoGlo substrate (Promega) in 25 mM Tris-HCl, 150 mM NaCl, pH 7.4 with 0.05% antifoam were added to each fraction and luminescence was read on a FlexStation 3 plate reader. The column was calibrated using the following molecular weight standards (Merck 69385-30MG): thyroglobulin from bovine thyroid (670 kDa), γ-globulins from bovine blood (150 kDa), ovalbumin (44.3 kDa), ribonuclease A type I-A (13.7 kDa), and p-aminobenzoic acid (137 Da), yielding a log-linear relationship between elution volume and molecular weight that was used to assign elution positions to oligomeric states.

### Alpha-Fold

Three-dimensional protein structures of apolipoprotein E isoforms (ApoE2, ApoE3, and ApoE4) with the SmBiT and LgBiT tags were predicted using the AlphaFold Server powered by AlphaFold 3(30, 31) and developed by Google DeepMind. The canonical full-length amino acid sequence of human APOE (P02649) was used as the reference sequence for ApoE3. Sequences were submitted to the AlphaFold Server individually and as a dimer with default parameters. Predicted structures were downloaded in PDB format and visualized using PyMol (Schrödinger).

### Statistical Analysis

All experiments were performed in a minimum of three independent biological replicates. Data are presented as mean ± standard error of the mean (SEM). For the self-association endpoint data, statistical comparisons were made using the Kruskal–Wallis test for overall group differences, followed by pairwise Mann–Whitney U tests with Holm correction for multiple comparisons. Significance thresholds were set at *p < 0.05, **p < 0.01, ***p < 0.001, ****p < 0.0001. Outliers were identified and excluded using the ROUT method (Q = 1%). Sensitivity analyses with and without excluded points did not alter the interpretation of the primary comparisons

## RESULTS

### Development of a Split-Luciferase ApoE self-association Assay

To quantify ApoE self-association without the need for protein isolation, we fused the SmBiT and LgBiT subunits of NanoLuc luciferase to the C-terminus or N-terminus of full-length ApoE2, ApoE3, and ApoE4 (Figure 1A). We then co-expressed these in HEK cells using transient transfection. When ApoE-SmBiT and ApoE-LgBiT dimerize, the SmBiT and LgBiT subunits are brought into proximity, reconstituting luciferase activity and producing a luminescent signal proportional to the extent of dimerization(26). Transfection with both SmBiT and LgBiT tagged versions of protein was necessary to generate luciferase signal (Figure 1B). We found that the signal generated was much greater for C-terminally tagged versions of the proteins when compared with N-terminally tagged versions (Figure 1C) and so proceeded to use C-terminally tagged protein throughout.

**Figure 1.**
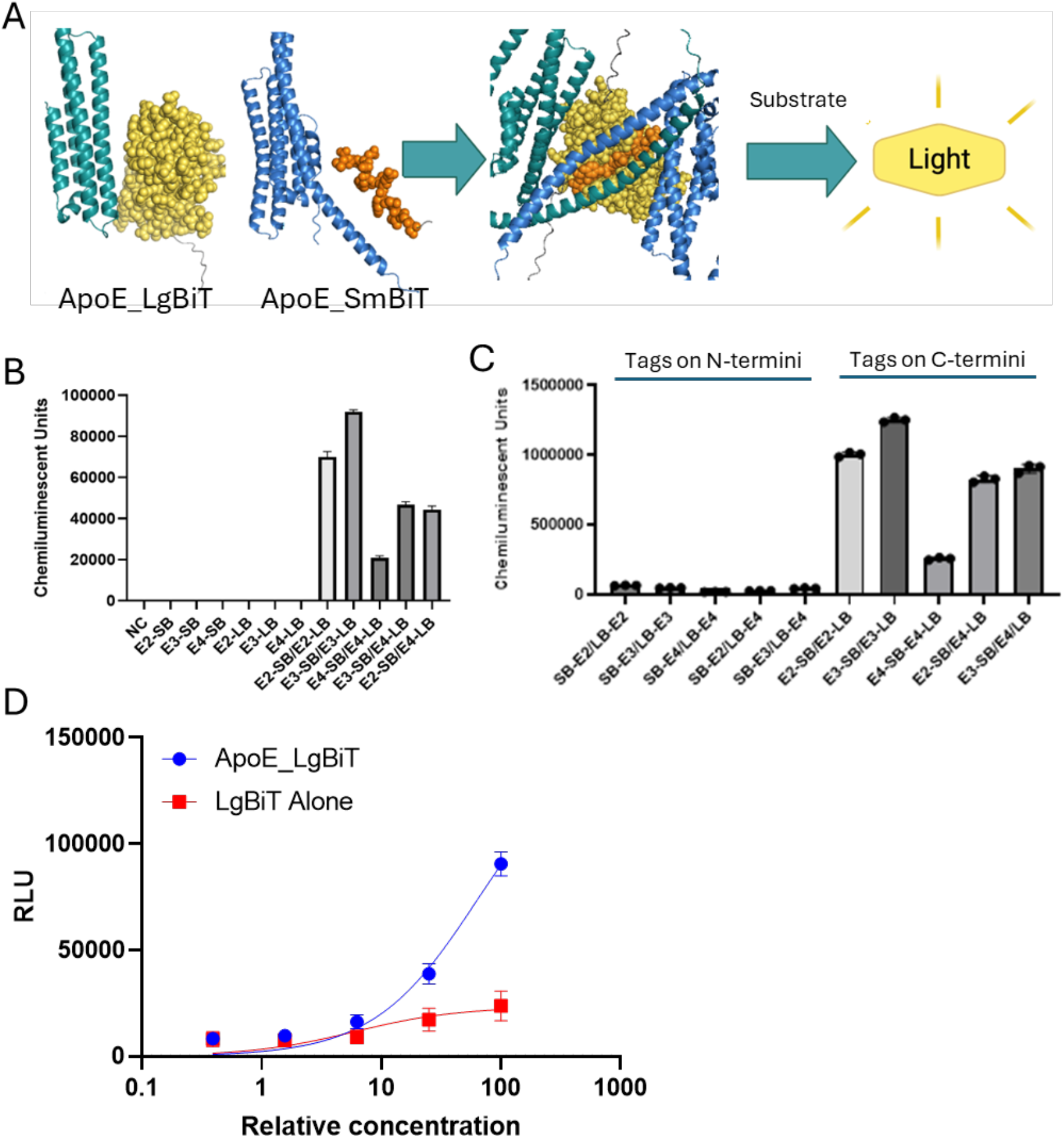
Schematic of the NanoBiT split-luciferase ApoE self-association assay. **(A)** ApoE-SmBiT and ApoE-LgBiT are co-secreted from transfected HEK293 cells. Self-association of ApoE brings the SmBiT and LgBiT subunits into proximity, reconstituting NanoLuc luciferase activity and producing a luminescent signal quantified as relative light units (RLU). A three-dimensional predicted model of the ApoE SmBiT–LgBiT complex was generated using AlphaFold3(30, 31). **(B)** Luminescence intensity from the co-transfection of HEK293 cells with different C-terminally tagged ApoE constructs **(C)** Luminescence intensity from the co-transfection of HEK293 cells with N and C-terminally tagged constructs. **(D)** ApoE3-LgBiT media or LgBiT alone was combined with ApoE3-SmBiT media and luminescence was measured across a 4-fold serial dilution series, with 100% representing the standard assay concentration. Error bars represent SEM. Luminescence is reported in arbitrary units (RLU.).

To confirm that the luminescence signal detected in the NanoBiT assay reflects a direct interaction between ApoE molecules rather than spontaneous complementation of the NanoBiT fragments themselves, we compared signal generated by ApoE3-LgBiT co-expressed with ApoE3-SmBiT against that generated by unfused LgBiT co-expressed with ApoE3-SmBiT. Across a 256-fold concentration range, ApoE3-LgBiT produced a concentration-dependent increase in luminescence that substantially exceeded the signal from LgBiT alone at all concentrations tested (Figure 1D). At the standard assay concentration (100%), ApoE3-LgBiT generated approximately 6-fold greater signal than LgBiT alone, demonstrating that the observed luminescence is driven primarily by ApoE–ApoE interaction rather than by non-specific or spontaneous NanoBiT complementation.

### ApoE4 Shows Enhanced Self-Association Relative to ApoE3 while C-terminal Variants Show Reduced Self-Association

To quantify the amount of ApoE self-association with normalized levels of ApoE across the different isoforms we chose to singly transfect ApoE-SmBiT or ApoE-LgBiT into HEK cells and collect the media. Following normalization of ApoE concentration using western blot we combined ApoE-SmBiT and ApoE-LgBiT as homodimers of the same isoform in an approximately 7:1 ratio SmBiT to LgBiT by protein concentration. This ensured the LgBiT tagged ApoE was the rate limiting step. AS samples were normalized to equal relative ApoE input across constructs on the basis of western blot band intensity and absolute ApoE concentrations were not determined. Accordingly, the assay provides a comparative self-association readout across matched relative-input conditions rather than a direct measure of binding affinity. We found that ApoE4 constructs produced significantly greater luminescence than ApoE3 (Mann-Whitney U, p < 0.0001, Holm-corrected; Figure 2), demonstrating that ApoE4 has significantly enhanced self-association relative to ApoE3. We found that ApoE2 was not different from ApoE3.

**Figure 2.**
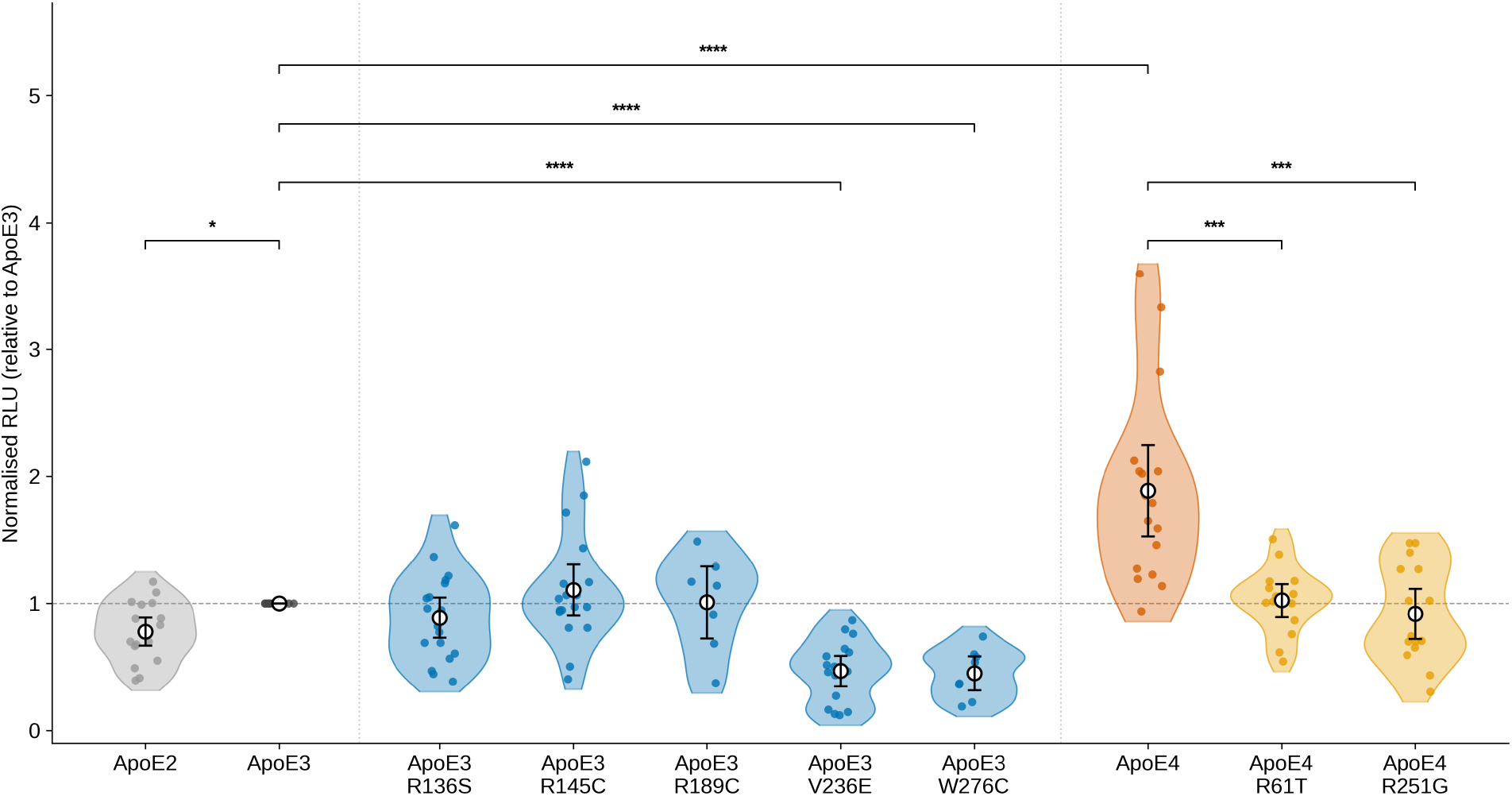
Impact of ApoE isoform and variant on self-association. Normalized maximum relative light units (RLU) from the endpoint self-association assay for each isoform and variant. Data are shown as violin plots with individual data points overlaid. ApoE3 is used as the reference (normalised to 1.0). Statistical comparisons: Kruskal–Wallis H = 71.4, p = 7.99 × 10^−12^; pairwise Mann–Whitney U with Holm correction. ****p < 0.0001, ***p < 0.001, *p < 0.05. RLU, relative light units.

We then used site-directed mutagenesis to generate SmBiT and LgBiT tagged versions of several rare variants of ApoE and tested these as homodimers using the same method. Two *APOE* ε*3*-backbone protective variants, Jacksonville (V236E)(19) and W276C(20), both produced significantly less luminescence than wild-type ApoE3 (Jacksonville: p < 0.0001; W276C: p < 0.0001; Holm-corrected Mann-Whitney U; Figure 2), reducing self-association well below ApoE3 levels

Similarly two variants on the *APOE ε4*-backbone, the naturally occurring protective variant R251G (19) and the predicted mutant R61T, both restored self-association to ApoE3-equivalent levels (R251G: p = ns vs ApoE3, p < 0.001 vs ApoE4; R61T: p = ns vs ApoE3, p < 0.001 vs ApoE4; Holm-corrected Mann-Whitney U; Figure 2). It is predicted that R61T disrupts the Arg61–Glu255 salt bridge responsible for ApoE4 domain interaction without altering the C-terminal sequence(11), and its normalisation of self-association to ApoE3 levels indicates that N-terminal domain interaction propagates allosterically to enhance C-terminal self-association in ApoE4.

Meanwhile the *APOE ε3*-backbone variants Christchurch (R136S)(21), R145C(22), and the Admixed American risk variant R189C (20)(OR = 3.35) did not significantly alter self-association relative to ApoE3 (all p = ns, Holm-corrected Mann-Whitney U; Figure 2).

### The self-association signal is predominantly driven by higher-order oligomers

Throughout this paper, we have used ‘self-association’ to refer to the NanoBiT complementation signal, which reflects the propensity of secreted, lipid-poor ApoE to form non-covalent complexes in conditioned medium. This signal does not distinguish between dimers, tetramers or higher-order oligomers, and is distinct from the cysteine-dependent disulfide-linked dimers described in plasma and CSF(32). To characterize the oligomeric state of secreted ApoE and to determine the distribution of ApoE-LgBiT across oligomeric species, we performed SEC on conditioned medium from HEK293 cells expressing ApoE3-LgBiT alone. ApoE3-LgBiT eluted predominantly in fractions corresponding to ∼66–106 kDa (Figure 3A), consistent with ApoE-LgBiT eluting as a dimer or a monomer.

**Figure 3.**
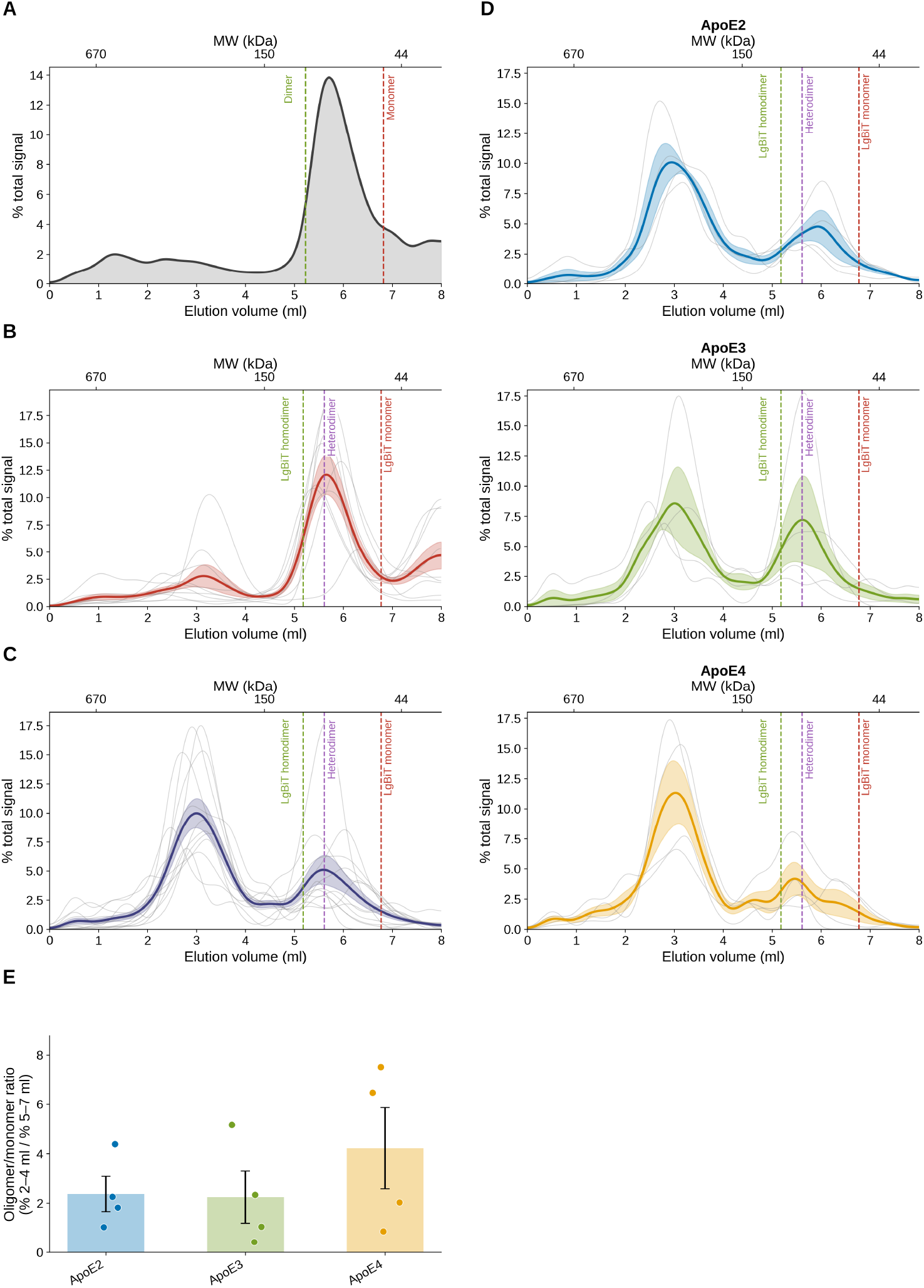
Size exclusion chromatography of ApoE-LgBiT reveals isoform-independent oligomeric distribution. **(A)** Elution profile of ApoE3-LgBiT alone (no SmBiT co-expression), detected by HiBiT lytic assay. Dashed lines indicate the expected elution volumes of the LgBiT monomer (53 kDa, red) and homodimer (104 kDa, green) based on the molecular weight calibration curve. **(B)** ApoE-LgBiT elution profile detected by HiBiT lytic assay across all genotypes. Individual runs are shown as grey traces. **(C)** SmBiT/LgBiT complementation signal across all genotypes. Grand mean ± SEM of all 12 runs (4 experiments × 3 genotypes). Individual runs are shown as grey traces. **(D)** Per-genotype SmBiT/LgBiT complementation profiles. Mean ± SEM for ApoE2, ApoE3, and ApoE4 (n = 4 independent experiments). Individual runs are shown as grey traces. **(E)** Oligomer/monomer ratio for each genotype, calculated as the percentage of total signal eluting between 2–4 ml divided by the percentage eluting between 5–7 ml, from the −HiBiT complementation signal. Bars show mean ± SEM; individual data points represent independent experiments (n = 4). One-way ANOVA: F = 0.86, p = 0.457.

We then applied HiBiT to the fractions from the SEC on mixed conditioned medium containing both ApoE-SmBiT and ApoE-LgBiT, and measured luciferase activity per fraction to detect the distribution of ApoE-LgBiT across the elution profile and found that it was similarly eluting in fractions consistent with homodimers of ApoE-LgBiT or heterodimers of ApoE-LgBiT/ApoE-SmBiT (Figure 3B). To determine the contribution of different oligomeric species to the overall signal from the assay, we measured luciferase activity per fraction after adding only substrate and found that a peak corresponding to ∼250–500 kDa (Figure 3C), substantially larger than the ApoE-LgBiT elution peak observed, indicating that ApoE-LgBiT is incorporated into higher-order ApoE-SmBiT oligomers in the mixed condition. Approximately one-third of the signal co-eluted with the monomeric/dimeric ApoE-LgBiT peak, consistent with true 1:1 heterodimers, while the remaining two-thirds co-eluted with higher-molecular-weight fractions. These profiles were similar across ApoE2, ApoE3, and ApoE4 (Figure 3D,E).

## DISCUSSION

Here we describe a quantitative, cell-based split-luciferase assay for ApoE self-association and use it to characterize the effects of common isoforms, naturally occurring disease-risk and protective variants, and an engineered structural probe on ApoE C-terminal self-association. We show that ApoE4 shows significantly greater self-association than ApoE3, and this enhanced self-association is mechanistically linked to N-terminal domain interaction as the R61T variant which is predicted to disrupt the Arg61–Glu255 salt bridge(11), restores signal to ApoE3-equivalent levels. Interestingly the protective *APOE ε4*-backbone variant R251G is also able to reduce the level of self-association to that of ApoE3.

Among the *APOE ε3*-backbone variants, Jacksonville (V236E) and W276C both reduce self-association below ApoE3 levels, which highlights their location within the C-terminal domain which has previously been shown to be important for ApoE oligomerization(8, 15, 19, 20). By contrast, the Christchurch variant (R136S), R145C, R189C, and ApoE2 do not significantly alter signal relative to ApoE3, indicating that their effects on AD risk are mediated through mechanisms independent of C-terminal self-association(20–22). Collectively, these data reveal that variants with the strongest impact on ApoE self-association are those which alter the C-terminal domain, while other variants modulate risk through structurally and functionally distinct pathways.

The enhanced self-association of ApoE4 relative to ApoE3 is consistent with prior structural and biophysical studies(14, 33–35). ApoE4 domain interaction is predicted to be altered compared with ApoE3 and is thought to promote intermolecular coiled-coil interactions although other mechanisms are also predicted(10, 25, 36). The observation that the mutant R61T, which is predicted to disrupt domain interaction without altering the C-terminal sequence, normalizes signal in this assay to ApoE3 levels provides mechanistic evidence that while the C-terminal domain of ApoE is undoubtedly crucial for ApoE oligomerization, the N-terminal domain interaction of ApoE4 impacts self-association(11, 12, 24, 25). This is consistent with molecular dynamics simulations showing that R61T increases dynamics in the hinge region connecting the two domains, propagating structural changes to distal functional regions of the protein(11), and with cellular studies demonstrating that ApoE4-R61T behaves like ApoE3 in terms of mitochondrial function, neurite outgrowth, and intracellular trafficking(37, 38).

The finding that the naturally occurring human variant R251G restores ApoE4 to ApoE3-like signal is particularly informative. R251G is located in the C-terminal domain at a position predicted to lie within the oligomerization interface(12), and the Arg-to-Gly substitution likely directly disrupts the intermolecular contacts that stabilize ApoE4 self-association. Human genetic data show that *APOE* ε4*R251G counterbalances the AD risk conferred by the ε4 allele(19), and our data provide a plausible structural mechanism. This interpretation is consistent with the broader model in which ApoE4 self-aggregation sequesters the protein in a lipid-poor, functionally impaired state(18, 24, 35).

The Jacksonville variant (V236E) reduced signal below ApoE3 levels in this assay, consistent with the mechanistic studies of Liu et al. (2021)(15), who demonstrated that V236E reduces ApoE aggregation and enhances lipidation in astrocyte-conditioned medium and human brain tissue. Val236 is located in the C-terminal amphipathic helix that forms the core of the intermolecular coiled-coil interface (10, 12), and the introduction of a charged glutamate residue at this position likely disrupts hydrophobic packing. Similarly W276C, which is strongly protective(20), significantly reduced signal below ApoE3 levels, mirroring the Jacksonville variant and suggesting that reduced C-terminal self-association may be a shared mechanism of protection for C-terminal variants. The fact that these variants reduce signal below ApoE3 levels suggests that even the baseline level of ApoE3 self-association may be suboptimal for lipid transport, and that further reduction in self-association, as conferred by V236E or W276C, may enhance the availability of monomeric, lipid-binding-competent ApoE.

In contrast, the Christchurch variant (R136S) did not alter self-association signal relative to ApoE3. The protective effects of Christchurch have been attributed to reduced HSPG binding, which limits ApoE-mediated tau uptake and propagation, and to suppression of microglial cGAS-STING signaling (21, 39, 40). Similarly, ApoE2 did not differ from ApoE3 in this assay, consistent with the view that ApoE2’s protective effects are mediated primarily through altered LDLR binding and amyloid-β clearance rather than altered self-association(41, 42). Neither of the *APOE* ε3 backbone increased risk variants R189C or R145C altered self-association. Our data support the conclusion that APOE2, Christchurch, R145C, and R189C protect against or increase AD risk through mechanisms orthogonal to C-terminal self-association, while Jacksonville, R251G, and W276C may act at least in part through this interface (Figure 2).

It is important to note that the self-association measured here is distinct from the cysteine-dependent disulfide-linked dimers of ApoE that have been described in plasma and CSF (32, 43). Those disulfide-linked species are unique to ApoE3 and ApoE2, which carry cysteine residues at position 112 and/or 158, and cannot form in ApoE4. These cysteine-dependent ApoE3 homodimers and ApoE3–apoA-II complexes have been linked to HDL-related behavior and redox buffering (44, 45), and plasma ApoE homodimer levels are reduced in APOEε3-carrying AD patients relative to controls(43), suggesting that these disulfide-linked species may themselves be functionally relevant in ways not captured by the NanoBiT assay. The NanoBiT signal instead reflects the non-covalent, C-terminal-domain-mediated self-association that is common to all three isoforms and has been characterized biophysically as a mixture of dimers, tetramers, and higher-order oligomers in equilibrium (14). This form of self-association is functionally significant because dissociation to monomer is thought to be required for high-affinity lipid binding(18) and may influence downstream receptor interactions, making the non-covalent oligomeric pool the relevant target for understanding isoform-dependent differences in ApoE lipidation and cholesterol transport. The SEC fractionation data provides important mechanistic context for interpreting the signal seen in this assay. The finding that ApoE-LgBiT elutes predominantly at ∼88–106 kDa across all three isoforms indicates that the LgBiT tag potentially prevents oligomerization and that the isoform differences in luminescence reflect genuine differences in self-association propensity of the ApoE-LgBiT to integrate into oligomers generated by the ApoE-SmBiT population. This is a limitation of large tags such as LgBiT (18kDa) and a strength of very small tags such as SmBiT (1.3 kDa).

Several limitations of the current study should be noted. First, the assay measures self-association of secreted, lipid-poor ApoE in conditioned medium, which may not fully recapitulate the behavior of lipidated ApoE in the CNS. ApoE lipidation is known to shift the monomer– oligomer equilibrium towards monomer (18, 24, 35), and the isoform differences reported here may therefore represent an upper bound on the magnitude of self-association differences in a fully lipidated context. Second, the main assay was performed using homodimer combinations of each isoform. The majority of *APOE ε4* carriers are heterozygous (ε3/ε4), and ApoE3/ApoE4 heterodimers have been shown to adopt conformations distinct from either homodimer(46). Further studies will directly address this by characterizing ApoE3/ApoE4 and ApoE3/Jacksonville heterodimeric combinations using the same assay. Third, the HEK293 expression system does not fully recapitulate the lipidation state or post-translational modifications of ApoE in primary astrocytes or neurons, and future work should validate key findings in more physiologically relevant cell types(13, 15, 47). Fourth, the ApoE-LgBiT appears to be forced to be a monomer or dimer which is not physiologically relevant. However, that is also a potential strength of the system as if the ApoE-LgBiT was similarly aggregated to the ApoE-SmBit then it is likely we would get no signal as both tags would be buried in ApoE aggregates unable to form new protein-protein interactions.

From a therapeutic perspective, the data presented here support the concept that reducing ApoE4 self-association is a viable strategy for correcting ApoE4 function. The mechanistic insight provided by R61T, that domain interaction drives enhanced C-terminal self-association, suggests that small-molecule structure correctors targeting this interaction may simultaneously normalize both ApoE4 conformation and oligomerization. Such correctors have already been shown to restore ApoE3-like behavior in iPSC-derived neurons(37, 48), and the NanoBiT self-association assay described here provides a quantitative, cell-based readout that could be adapted for high-throughput screening of compounds that modulate ApoE self-association. The finding that W276C reduces self-association to a similar extent to Jacksonville further validates the C-terminal domain as a druggable target.

## CONCLUSION

We have developed a sensitive, quantitative split-luciferase assay for ApoE self-association in serum-free conditioned medium from transfected HEK293 cells. ApoE4 shows significantly enhanced self-association relative to ApoE3, driven by the ApoE4 C-terminal domain interaction. Two APOE3-backbone C-terminal variants, Jacksonville (V236E) and W276C, reduce ApoE3 self-association below wild-type levels while the APOE4-backbone protective variant R251G and the predicted domain-interaction mutant R61T each reduce ApoE4 self-association to ApoE3-equivalent levels. Variants outside the C-terminal domain, Christchurch (R136S), R145C, and R189C, do not alter self-association, indicating that these variants modulate AD risk through orthogonal mechanisms. Together, these data establish the C-terminal self-association interface as an element of isoform-specific ApoE behavior and support its targeting as a therapeutic strategy for ApoE4-driven AD risk.

## Data Availability

All data produced in the present study are available upon reasonable request to the authors

## Acknowledgements and funding

The authors acknowledge funding from Alzheimer’s Drug Discovery Foundation (ADDF) to R. Jackson, from Alzheimer’s San Diego to M. Jackson and grants from the National Institutes of Health (NIH) 5R56AG080525 (M. Jackson and B. Hyman), and 1R01AG086914 (M. Jackson). We would like to acknowledge funding from the JPB Foundation, and to the Mass ADRC (P30AG062421). R Jackson would also like to acknowledge the help, support and guidance from the Harrington Discovery Institute.

---

**Supplementary Table 1.**
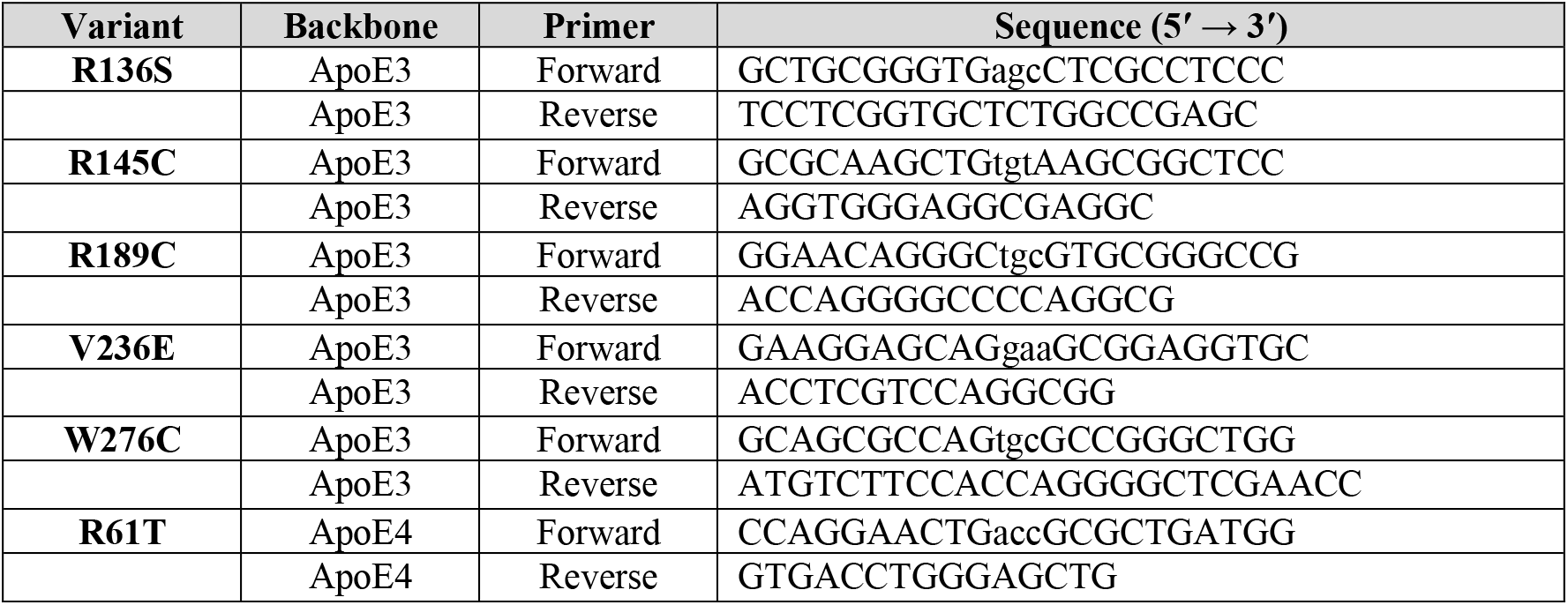
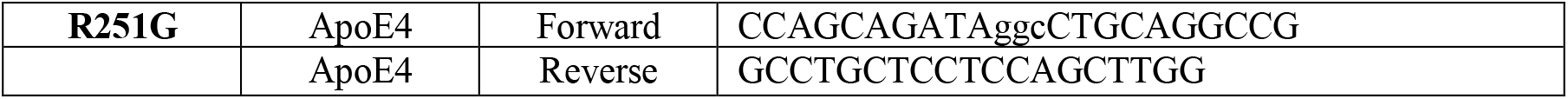
Oligonucleotide primers used for site-directed mutagenesis of APOE variants. Mutagenic nucleotides are shown in lowercase. All primers were designed using the NEBaseChanger® Primer Design Tool (New England Biolabs) and used with the Q5® Site-Directed Mutagenesis Kit (New England Biolabs). Variants were introduced into the indicated ApoE backbone.

**Supplementary Figure 1.**
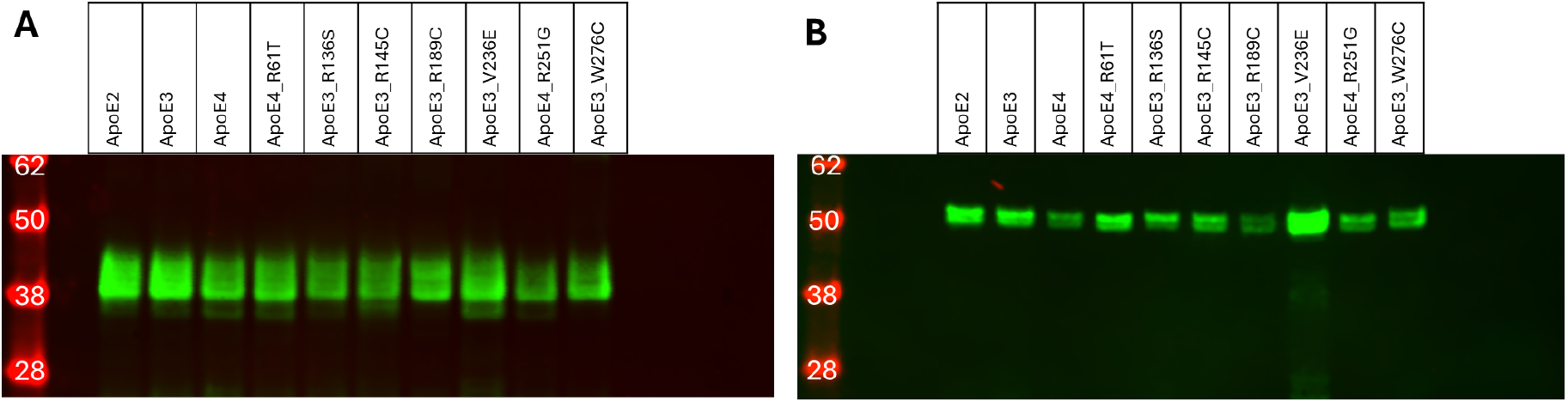
Expression of ApoE-NanoBiT fusion proteins in HEK293T cells. Western blot showing expression of ApoE-SmBiT (A) and ApoE-LgBiT (B) constructs in whole-cell lysates from transfected HEK293T cells. Each lane represents an independent biological replicate. Blots were probed with an anti-ApoE antibody. Molecular weight markers (kDa) are indicated. Expected band sizes reflect the molecular weight of ApoE (∼37 kDa) plus the SmBiT (1.3 kDa) or LgBiT (17.6 kDa) tag.

**Supplementary Figure 2.**
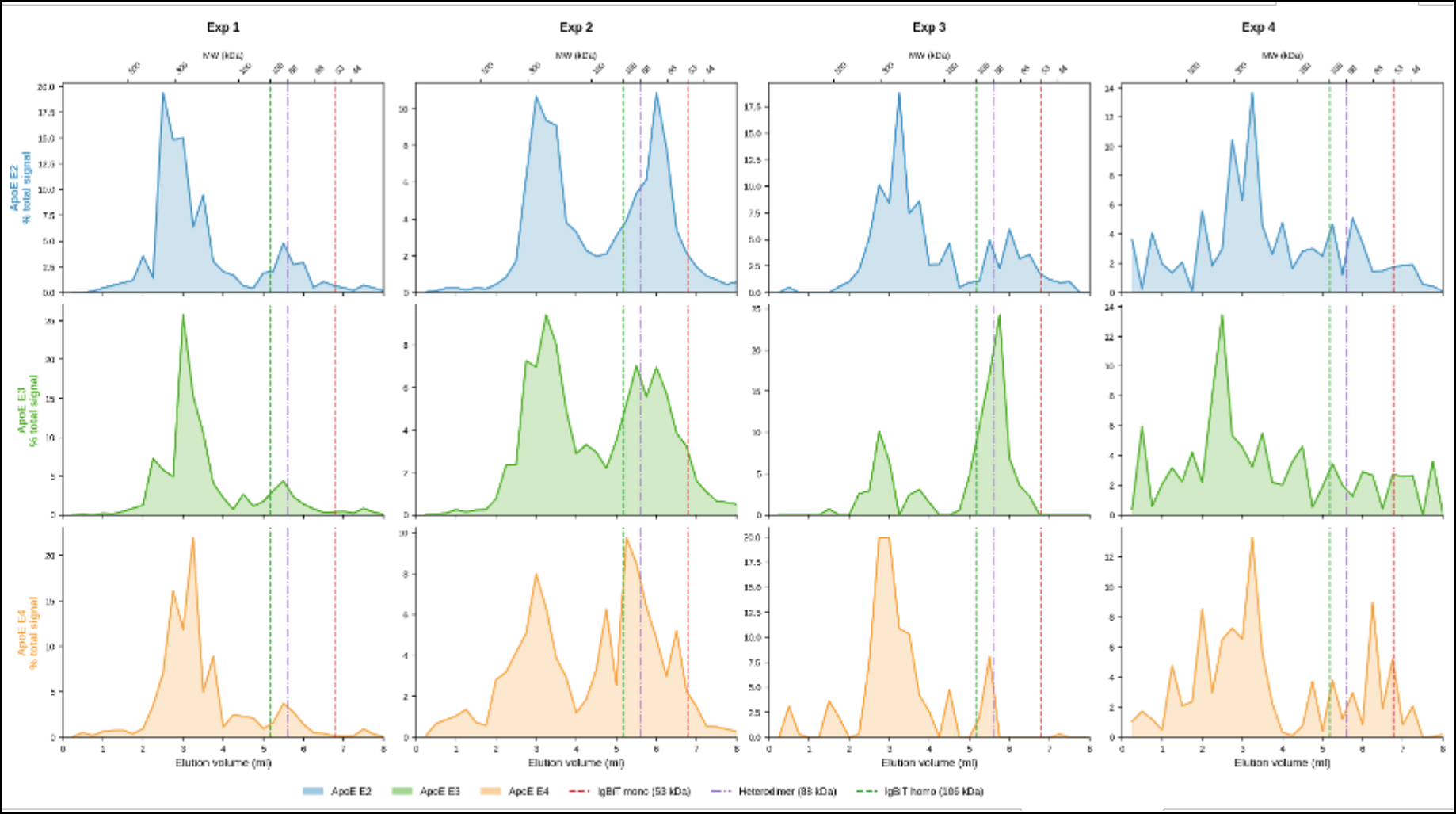
Individual SEC traces for all ApoE-NanoBiT constructs. Raw size-exclusion chromatography (SEC) for each individual replicate of all ApoE-NanoBiT constructs analyzed in this study. Each line represents a single chromatographic run; no smoothing, averaging, or baseline correction has been applied. Traces are grouped by construct and colored as in Figure 3. The elution positions of molecular weight standards are indicated by dashed vertical lines. Individual replicate traces are shown here to support the mean traces presented in Figure 3 and to allow full assessment of run-to-run variability.

